# Limited Capacity for Ipsilateral Secondary Motor Areas to Support Hand Function Post-Stroke

**DOI:** 10.1101/19012336

**Authors:** Kevin B. Wilkins, Jun Yao, Meriel Owen, Haleh Karbasforoushan, Carolina Carmona, Julius P.A. Dewald

## Abstract

Recent findings have shown connections of ipsilateral cortico-reticulospinal tract (CRST), predominantly originating from secondary motor areas, to not only proximal but also distal portions of the arm. In unilateral stroke, CRST from the ipsilateral side is intact and thus has been proposed as a possible backup system for post-stroke rehabilitation even for the hand. We argue that although CRST from ipsilateral secondary motor areas can provide control for proximal joints, it is insufficient to control either hand or coordinated shoulder and hand movements due to its extensive branching compared to contralateral corticospinal tract. To address this issue, we combined MRI, high-density EEG, and robotics in 17 individuals with severe chronic hemiparetic stroke and 12 age-matched controls. We tested for changes in structural morphometry of the sensorimotor cortex and found that individuals with stroke demonstrated higher gray matter density in secondary motor areas ipsilateral to the paretic arm compared to controls. We then measured cortical activity while participants attempted to generate hand opening either supported on a table or while lifting against a shoulder abduction load. The addition of shoulder abduction during hand opening increased reliance on ipsilateral secondary motor areas in stroke, but not controls. Crucially, increased use of ipsilateral secondary motor areas was associated with decreased hand opening ability while lifting the arm due to involuntary coupling between the shoulder and wrist/finger flexors. Together, this evidence implicates a compensatory role for ipsilateral (i.e., contralesional) secondary motor areas post-stroke, but with limited capacity to support hand function.

## I. Introduction

Hand function is often significantly impacted post-stroke, particularly in individuals with moderate to severe motor impairments. This is partially attributed to damage to the corticospinal tract (CST), which is the primary motor tract controlling the hand in healthy individuals^1,2^. Following damage to the CST, individuals with stroke demonstrate increased reliance on the ipsilateral (i.e., contralesional) sensorimotor cortices when attempting to move the paretic arm^3,4^. This may reflect recruitment of uncrossed indirect motor pathways originating from ipsilateral sensorimotor cortices to generate motor output^5^. However, the question remains whether the ipsilateral sensorimotor cortex and associated alternate motor pathways have the capacity to support hand function.

Uncrossed cortico-bulbospinal fibers originating from the cortex ipsilateral to the moving arm, such as the corticoreticulospinal tract (CRST), may serve as a compensatory backup system to control the paretic arm following damage to CST^5,6^. One piece of evidence in support of this possibility of post-stroke reliance on CRST comes from the finding that following a pyramidal CST lesion in monkeys, connections between the ipsilateral reticular formation and paretic wrist flexors are strengthened^7^. Importantly, these ipsilateral tracts originate predominantly from secondary motor regions such as the supplementary motor area (SMA) and premotor cortex (PM) compared to CST which predominantly originates from primary motor cortex (M1)^8-11^. Although the innervations from these tracts originating from secondary motor areas were thought to be limited to trunk and proximal muscles^11,12^, more recent work demonstrates that they also innervate distal muscles such as the wrist and finger flexors^13,14^. This raises the possibility that they could be involved in subsequent hand recovery following stroke^15^. However, the reticulospinal tract branches more extensively at the spinal cord compared to the CST^15-18^. Consequently, these pathways are not able to selectively activate individual muscles in the manner of CST and may not be sufficient for dexterous hand control^19^.

One of the main points of evidence for compensatory use of these uncrossed cortico-bulbospinal pathways following stroke in humans is the presence of abnormal coupling between the shoulder and the rest of the arm and hand. Lifting at the shoulder leads to abnormal coupling between shoulder abductors and elbow/forearm and finger flexor muscles that reduces reaching distance and hand opening ability, termed the flexion synergy^20-23^. In fact, lifting at the shoulder can lead to involuntary closing during attempted opening in individuals with more severe impairments^23^. Whereas the damage to CST accounts for the weakness, or inability to fully activate muscles^24,25^, it does not affect the presence of this abnormal coupling. Meanwhile, increased use of ipsilateral cortico-bulbospinal pathways originating from secondary motor areas could account for this coupling due to its more extensive branching at the spinal cord. This extensive branching limits the ability for these tracts to individually activate muscles, and instead leads to activation of multiple muscle groups simultaneously. Additionally, these pathways innervate a greater proportion of flexor muscles compared to extensors, and stimulation of these pathways preferentially elicits EMG activity in ipsilateral flexor muscles in the monkey^12,26^. Therefore, attempting to drive movement of the arm via these compensatory ipsilateral pathways originating from secondary motor areas may allow individuals to generate greater activity at the more proximal portions of the arm, but at the detriment of individual joint control and distal hand function, especially hand opening.

The goal of the current study was to investigate the potential compensatory role of ipsilateral secondary motor regions post-stroke and evaluate their capacity to support hand function following damage to CST. We hypothesized that individuals with a hemiparetic stroke would increasingly rely on ipsilateral secondary motor areas as compensation for damage to the lesioned hemisphere as the demand of the task increased, but that increased use of these areas would reduce hand opening ability due to the flexion synergy. To test this hypothesis, we first assessed any long-term structural changes in ipsilateral cortex using magnetic resonance imaging (MRI). Then, cortical activity was measured using high density electroencephalography (EEG) with measures of motor performance in a robotic controlled environment to link cortical activity to behavior. Cortical activity was measured during two tasks: 1. Hand opening in isolation and 2. Hand opening in conjunction with shoulder abduction (i.e., lifting). Cortical activity was compared with hand performance during these two conditions. We specifically examined grasping pressure as an indicator for hand opening ability since individuals with severe motor impairment are not able to open their hand and instead demonstrate involuntary grasping pressure when attempting to open^23^. Thus, more involuntary grasping was an indicator of worse performance. We found that i) individuals with stroke demonstrated increased gray matter density within secondary motor areas ipsilateral to the paretic arm (i.e., contralesional sensorimotor cortex) compared to controls; ii) the addition of shoulder abduction during attempted hand opening increased reliance on ipsilateral secondary motor areas in stroke, but not controls; iii) increased use of the ipsilateral secondary motor areas was associated with greater involuntary grasping (i.e., reduced hand opening ability) due to the flexion synergy. Together, these results implicate an increased reliance on ipsilateral secondary motor areas and presumably ipsilateral cortico-bulbospinal tracts as a compensatory means to generate more shoulder abduction torque in the paretic arm post-stroke, but with limited capacity to support distal hand opening.

## II. Materials and Methods

### 2.1 Participant

Seventeen individuals with chronic hemiparetic stroke (mean age: 58.9 ± 7.6 yrs.) and moderate to severe impairment (Upper Extremity Fugl Meyer Assessment [UEFMA]: 10-38; mean = 20.8 ± 8.4) and twelve age-matched controls (mean age: 59.8 ± 7.7 yrs.) participated in this study. Demographic information for each participant is provided in Table 1 and lesion locations in Figure 1. All individuals with stroke were screened for inclusion by a licensed physical therapist. Inclusion criteria included being at least one year post-stroke, an UEFMA no greater than 40 out of 66, MRI compatibility, and subcortical lesions not extending into sensorimotor cortices. This study was approved by the Northwestern institutional review board and all participants gave written informed consent.

**Table 1.** Participant Demographics.

| Controls |  |  |  | Stroke |  |  |  |  |  |
| --- | --- | --- | --- | --- | --- | --- | --- | --- | --- |
| Participant | Age | Sex | Dominant Arm | Participant | Age | Sex | UEFMA | Years post stroke | Lesioned Hemisphere |
| P<sub>1</sub> | 60 | M | R | P<sub>1</sub> | 62 | F | 23 | 7 | L |
| P<sub>2</sub> | 59 | M | R | P<sub>2</sub> | 49 | M | 11 | 18 | L |
| P<sub>3</sub> | 45 | F | R | P<sub>3</sub> | 60 | M | 11 | 6 | R |
| P<sub>4</sub> | 74 | M | R | P<sub>4</sub> | 60 | M | 10 | 19 | R |
| P<sub>5</sub> | 68 | F | R | P<sub>5</sub> | 60 | M | 19 | 9 | R |
| P<sub>6</sub> | 61 | M | R | P<sub>6</sub> | 63 | M | 22 | 9 | L |
| P<sub>7</sub> | 61 | M | R | P<sub>7</sub> | 68 | M | 13 | 21 | L |
| P<sub>8</sub> | 48 | F | R | P<sub>8</sub> | 57 | M | 24 | 5 | L |
| P<sub>9</sub> | 60 | F | R | P<sub>9</sub> | 60 | F | 24 | 12 | R |
| P<sub>10</sub> | 54 | F | R | P<sub>10</sub> | 66 | M | 17 | 9 | R |
| P<sub>11</sub> | 61 | M | R | P<sub>11</sub> | 71 | M | 15 | 13 | R |
| P<sub>12</sub> | 66 | M | R | P<sub>12</sub> | 47 | M | 38 | 7 | R |
|  |  |  |  | P<sub>13</sub> | 65 | F | 16 | 31 | L |
|  |  |  |  | P<sub>14</sub> | 47 | M | 16 | 10 | R |
|  |  |  |  | P<sub>15</sub> | 44 | M | 38 | 4 | L |
|  |  |  |  | P<sub>16</sub> | 64 | M | 30 | 7 | L |
|  |  |  |  | P<sub>17</sub> | 58 | M | 26 | 3 | L |
| Average | 59.8 |  |  |  | 58.9 |  | 20.8 | 11.2 |  |
| ± | ± | | | | ± | | ± | ± | |
| Std | 7.7 |  |  |  | 7.6 |  | 8.4 | 7.1 |  |

UEFMA: Upper extremity Fugl-Meyer Assessment; Std: Standard Deviation

**Figure 1.**
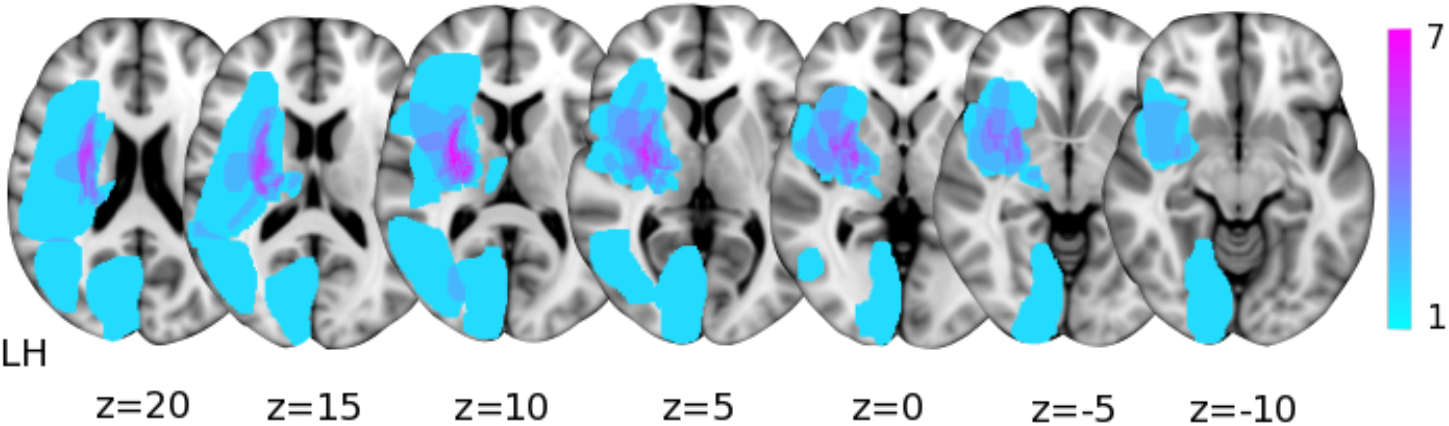
Subcortical lesion locations for the seventeen individuals with stroke overlaid on axial Montreal Neurological Institute T1 slices. The color bar indicates the number of participants with lesioned tissue in a particular voxel. LH indicates the lesioned hemisphere.

### 2.2 Experimental Protocols

#### 2.2.1. Structural Imaging of the Brain

Individuals participated in MRI scans at Northwestern University’s Center for Translation Imaging on a 3 Tesla Siemens Prisma scanner with a 64-channel head coil. Structural T1-weighted scans were acquired using an MP-RAGE sequence (TR=2.3s, TE=2.94ms, FOV 256×256mm^2^) producing an isotropic voxel resolution of 1×1×1 mm. Visual inspection of acquired images was performed immediately following the data acquisition to check the quality of the collected images and confirm stable head position.

#### 2.2.2. Functional Imaging related to hand and arm

In a separate experiment, functional imaging related to hand opening with or without arm lifting was examined using EEG. During the EEG experiment, participants sat in a Biodex chair (Biodex Medical Systems, Shirley, NY), which restrained the trunk with straps crossing the chest and abdomen. The participant’s paretic arm for individuals with stroke or dominant arm for healthy individuals was placed in a forearm-hand orthosis attached to the end effector of an admittance controlled robotic device (ACT^3D^) instrumented with a six degree of freedom (DOF) load cell (JR^3^ Inc., Woodland, CA).

At the beginning of each trial, participants moved their hand to a home position, with the shoulder at 85° abduction, 40° flexion, and the elbow at 90° flexion angle. The participant then received an auditory cue. Following the cue, participants relaxed at the home position for 5-7 s and then self-initiated either 1) a maximum attempted hand opening with the arm resting on a haptic table, or 2) a maximum attempted hand opening while lifting against 50% of maximum shoulder abduction torque (SABD50). Participants were instructed to avoid eye movements by focusing on a point and avoid movements of other body parts during the performance of each trial, which was visually confirmed by the experimenter. Participants performed 60-70 trials of each task, broken into blocks (one block consisted of 20-30 trials for a particular task). Rest periods varied between 15 to 60 seconds between trials and 10 minutes between blocks.

Scalp recordings were made with a 160-channel High-Density EEG system using active electrodes (Biosemi, Inc, Active II, Amsterdam, The Netherlands) mounted on a stretchable fabric cap based on a 10/20 system. The centers of all the electrode holders were attached with reflective markers. Simultaneously, EMGs were recorded from the extensor digitorum communis, flexor carpi radialis, and intermediate deltoid of the tested arm to assess timing of movement onset. All data were sampled at 2048 Hz. The impedance was kept below 50 kΩ for the duration of the experiment. The positions of EEG electrodes on the participant’s scalp were recorded with respect to a coordinate system defined by the nasion and pre-auricular notches using a Polaris Krios handheld scanner (NDI, Ontario, Canada). This allowed for coregistration of EEG electrodes with each participant’s anatomical MRI data. Additionally, for individuals with a stroke, involuntary grasping pressure during the two tasks was measured by a custom pressure sensor mat (Pressure Profile System Inc., CA) that was wrapped around a cylinder where the participant’s fingers/palm were placed around (see Figure 3A). Although participants were instructed to open their hand, individuals with severe chronic stroke cannot physically open their hand and instead display involuntary grasping due to the combination of weakness of finger extensor muscles and involuntary coactivation of finger flexor muscles^23^. Therefore, instead of directly measuring hand opening ability/aperture, grasping pressure was measured and used as a marker for inability to open the hand, with increased involuntary grasping pressure reflecting reduced hand opening ability. At the start of the experiment, maximum grasping forces were measured for the paretic hand, which were used for normalization purposes in the data analysis.

### 2.3 Data Analysis

#### 2.3.1. Structural Changes in Gray Matter Density

Anatomical T1 data were analyzed with FSL voxel-based morphometry (VBM) 1.1 (https://fsl.fmrib.ox.ac.uk/fsl/fslwiki/FSLVBM; Oxford University, Oxford, United Kingdom)^27^ using FSL tools^28^. T1 images for individuals with right hemisphere lesions were flipped to ensure that the lesions of all stroke participants were in the left hemisphere. The T1 images were then brain-extracted using the Brain Extraction Tool and segmented into gray matter using FAST4. The resulting gray matter partial volume images were aligned to Montreal Neurological Institute (MNI) 152 standard space using the affine registration tool FLIRT and averaged to create a study-specific gray matter template. Subsequently, individual gray matter partial volume images in native space were non-linearly registered to this template using FNIRT, modulated to correct for local expansion or contraction due to the non-linear component of the spatial transformation, and then smoothed with an isotropic Gaussian kernel with a sigma of 3 mm. These gray matter images were masked to only include the ipsilateral sensorimotor cortex including primary motor cortex, supplementary motor area, premotor cortex, and primary somatosensory cortex from the Human Motor Area Template^29^.

#### 2.3.2. Involuntary Grasping Pressure

The grasping pressure was calculated as the sum of max pressure generated by the I-IV digits during a given trial^23^ (see an example of the pressure generated in Figure 3B). Ensemble-averaged grasping pressure for each condition was then normalized by the maximum grasping pressure, which was calculated as the average of the largest 3 total grasping pressures during the max closing trials. Grasping pressure is thus referred to as the percent of pressure during a specific task compared to the individuals max closing pressure.

#### 2.3.3 Cortical activity related to hand opening and hand opening while lifting against load

EEG data were low pass filtered at 50 Hz, aligned to the earliest EMG onset of the 3 muscles, and segmented from −2200 to +200 ms (with EMG onset at 0 ms) using Brain Vision Analyzer 2 software (Brain Products, Gilching, Germany). Data were then visually inspected for the presence of artifacts. Trials exhibiting artifacts (e.g., eye blinks) were eliminated from further analysis. The remaining EEG trials were baseline-corrected (from −2180 to −2050 ms) and ensemble-averaged. The averaged EEG signals were down-sampled to 256 Hz and imported into CURRY 6 (Compumedics Neuroscan Ltd., El Paso, TX). The cortical current density strength (μA/mm^2^) in the time between 150 ms and 100 ms prior to EMG onset was computed using the standardized low resolution electromagnetic brain tomography (sLORETA) method (Lp = 1) based on a participant-specific boundary element method model with the regulation parameter automatically adjusted to achieve more than 99% variance accounted^30,31^. Possible sources were located on a cortical layer with 3 mm distance between each node. Although the inverse calculation was performed over the whole cortex, only the activity in bilateral sensorimotor cortices was further analyzed. Specific regions of interest (ROIs) included bilateral primary sensorimotor cortices (primary motor cortex (M1) + primary sensory cortex (S1)) and secondary motor cortices (supplementary motor area (SMA) + premotor area (PM)).

We used the estimated current density strengths to calculate a Laterality Index (LI = (C-I)/(C+I)), where C and I are the current density strengths from the contralateral and ipsilateral sensorimotor cortices relative to the moving hand/arm (i.e., combined primary sensorimotor and secondary motor cortices), respectively. LI reflects the relative contributions of contralateral versus ipsilateral sensorimotor cortices to the source activity, with a value close to +1 for a contralateral source distribution and −1 for an ipsilateral source distribution.

Additionally, we quantified a cortical activity ratio 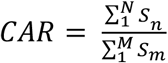 for each of the 4 ROIs, where *S*_*n*_ represents the current density strength of the *n*^*th*^ node, and N and M represent the number of nodes in one of the ROIs and the whole sensorimotor cortices, respectively. The cortical activity ratio reflects the relative strength from one ROI as normalized by the total combined strength of the 4 ROIs. When a significant effect of task in CAR was found, we further examined between-task difference in the sum of absolute amplitude activity in each ROI. This is to justify the possible interdependencies between regions (e.g., one region increasing in CAR can lead to a decrease in CAR in another even if the absolute activity does not change in the second region). However, measure of absolute activity can only be used for within-subject comparisons, due to the between subject variance in signal to noise ratio, scalp conductance, electrode impedance, etc.

### 2.4 Statistical Analysis

Statistics for the GM density were computed within FSL. A voxel-wise General Linear Model was applied with a Threshold-Free Cluster Enhancement^32^ to detect differences in gray matter density between individuals with stroke and controls. A voxel-based threshold of changes in gray matter density was set at *p* < 0.05 (Family-Wise Error Corrected; FWE). Statistics for the behavior and EEG were performed using SPSS (IBM, V23). A paired t-test was performed to assess any impact on task on the normalized grasping pressure in individuals with stroke. A 2 (group) x 2 (task) ANOVA was performed on LI for the EEG analysis. A 2 (group) x 2 (task) x 4 (region) ANOVA was performed on CAR for the EEG analysis. We performed post-hoc paired t-tests for any significant within-subject effect in ANOVA interactions. Pearson correlations were performed between significant cortical activity findings and grasping pressure. A *p* value of 0.05 or less was considered significant.

## III. Results

### 3.1 Differences in Gray Matter Density in Ipsilateral Sensorimotor Cortex

Structural differences in gray matter (GM) density within sensorimotor cortices were compared between individuals with stroke and healthy controls. Individuals with stroke demonstrated significant greater GM density compared to controls in two ipsilateral clusters: 1) a cluster residing in premotor cortex (peak voxel: x = 46, y = 6, z = 50, *t*-value = 5.17, *p* < 0.05 FWE corrected; Figure 2A), and 2) a cluster residing in primary somatosensory cortex (peak voxel: x = 48, y = −26, z = 58, *t*-value = 5.55, *p* < 0.05 FWE corrected; Figure 2B). Meanwhile, there were no regions that exhibited significantly greater GM density in controls compared to individuals with stroke within the ipsilateral sensorimotor cortex.

**Figure 2.**
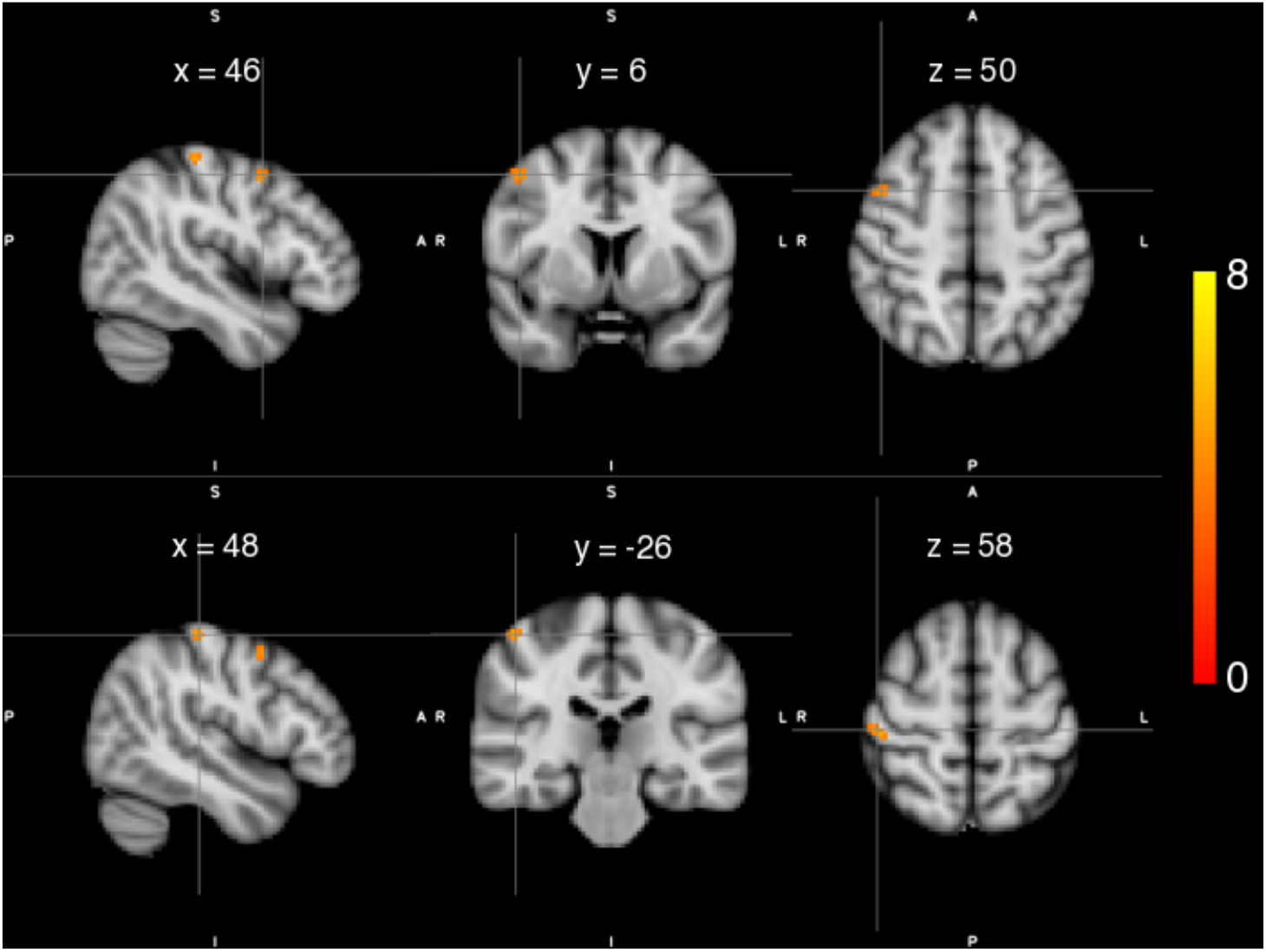
Statistical maps of gray matter (GM) density differences for individuals with stroke compared to healthy controls. Significantly higher GM density was observed in ipsilateral premotor cortex (Top) and ipsilateral primary somatosensory cortex (Bottom) in individuals with stroke compared to controls. Color maps indicate the thresholded t values at each voxel. A statistical threshold was set equivalent to *p* < 0.05 FWE.

### 3.2 Impact of Shoulder Abduction on Hand Opening Ability Post Stroke

Since the cohort of this study was primarily severely impaired and could not open their paretic hand, we measured grasping pressure as an indicator of inability to open the hand. We found that twelve of the seventeen individuals with chronic stroke could not open their hand off the cylinder and therefore included them in the grasping pressure analysis. An example for one individual’s grasping pressure during the two conditions is depicted in Figure 3C. Overall, these individuals demonstrated a significant increase in involuntary grasping pressure (reduced hand opening ability) with the addition of the SABD load compared to attempted hand opening on the table (*t*(11) = 3.16, *p* = 0.009; Figure 3D). Controls were not analyzed since they do not produce any involuntary grasping pressure during either condition, and a 50% max SABD load does not reduce hand opening ability^23^.

**Figure 3.**
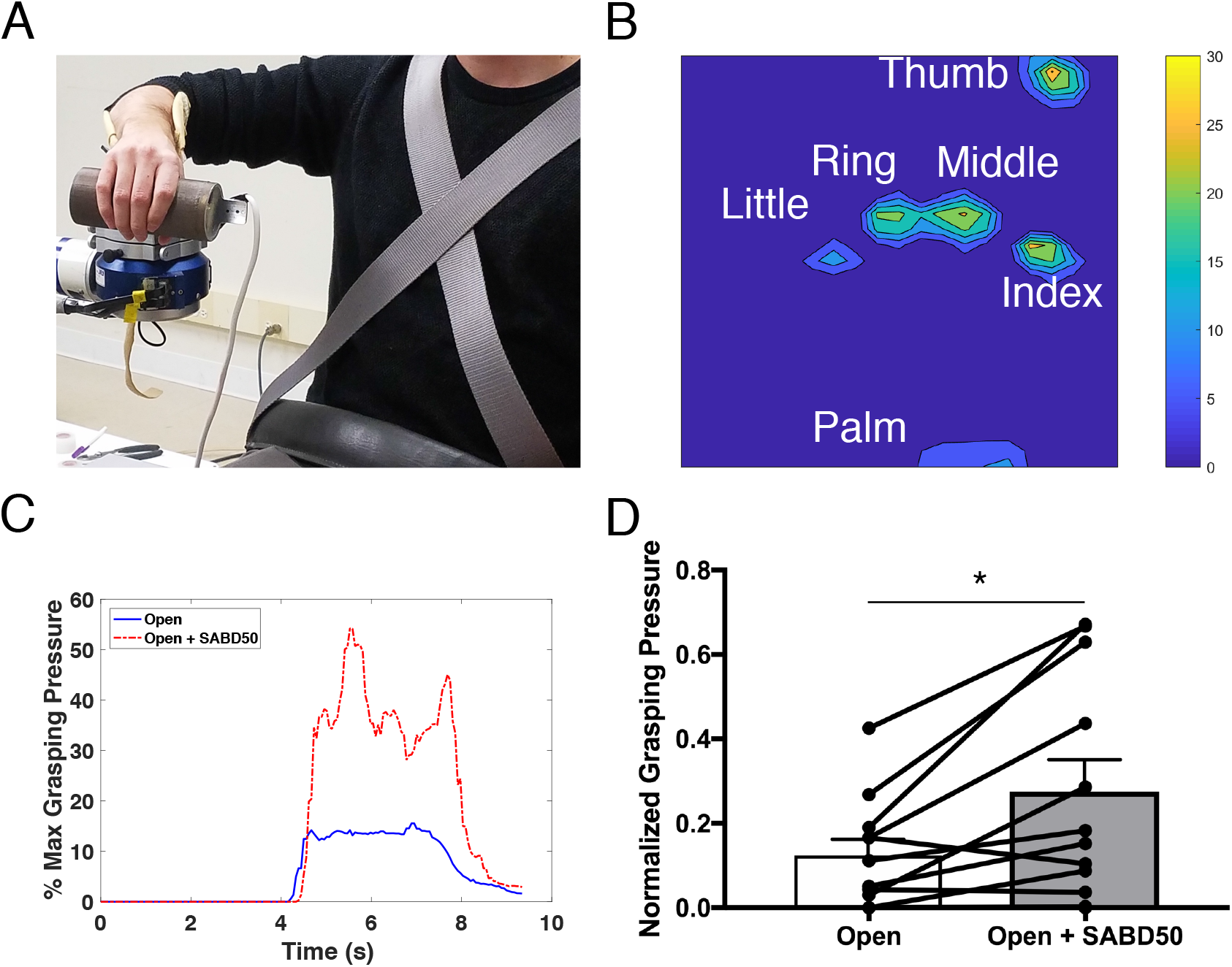
Shoulder abduction reduces hand opening ability in individuals with stroke. (A) The ACT-3D system with the attached forearm-hand orthosis equipped with a TactArray sensor mat to measure grasping pressure. (B) An example of grasping pressure measured by the TactArray sensor mat. (C) An example from one individual of grasping pressure over time for attempted hand opening on the table (solid Blue) and attempted hand opening while lifting against 50% max shoulder abduction (broken Red) depicted as the percentage of max grasping pressure. (D) Group averages with individual data overlaid of normalized grasping pressure for opening on the table vs. opening while lifting against 50% max shoulder abduction. Error bars depict SEM. * p < 0.05.

### 3.3 Impact of Shoulder Abduction on Cortical Activity

A 2 (group) x 2 (task) ANOVA was conducted to examine the effect of group and task on the laterality index (LI). There was a statistically significant interaction between the effects of group and task on LI (*F*(1,54) = 6.62, *p* = 0.013; Figure 4). Post hoc paired t-tests showed that LI was significantly lower (more ipsilateral) during the Open + SABD50 condition compared to opening on the table for individuals with stroke (*t*(16) = 3.16, *p* = 0.006). Meanwhile, controls showed no difference in LI between the two tasks.

**Figure 4.**
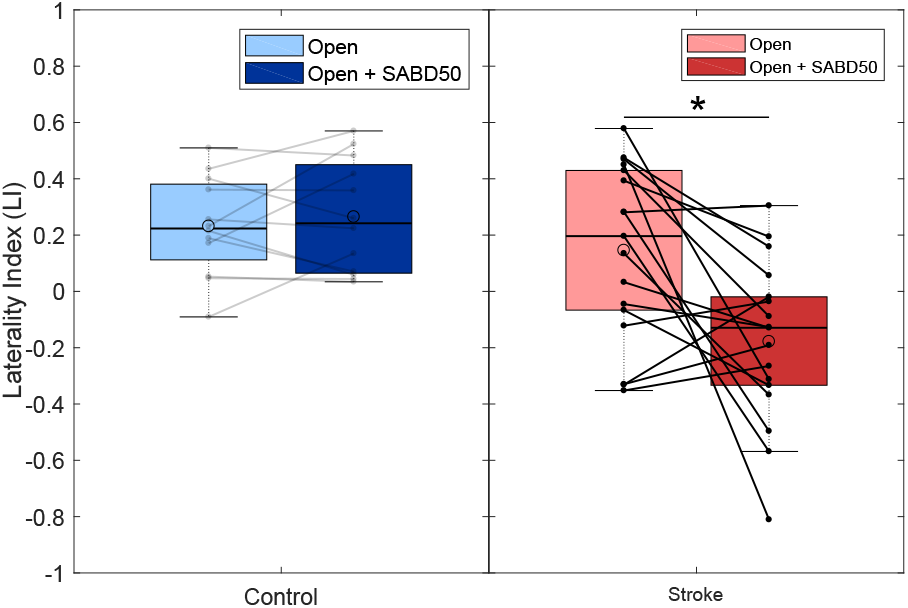
Shoulder abduction increases reliance on the ipsilateral hemisphere in stroke but not controls. Boxplots with individual data overlaid showing laterality index (LI) for controls (left; blue) and stroke (right; red) for hand opening on the table (light) and hand opening while lifting against 50% max shoulder abduction (dark). Controls show no difference between conditions, while the addition of SABD increases reliance on the ipsilateral hemisphere in individuals with stroke (i.e., negative LI). * p < 0.05.

A 2 (group) x 2 (task) x 4 (region) ANOVA was conducted to examine the effect of group, task, and region on CAR. There was a statistically significant three-way interaction between the effects of group, task, and region on CAR (*F*(3,216) = 3.01, *p* = 0.03; Figure 5). Post hoc paired t-tests showed that in individuals with stroke, the addition of lifting to opening caused significantly increased CAR in ipsilateral secondary motor areas (i-SMA/PM, *t*(16)=3.01, *p*=0.008) and decreased CAR in contralateral primary sensorimotor cortices (c-M1/S1, *t*(16)=2.73, *p*=0.015). In controls, there were no differences between any of the regions during the two tasks.

**Figure 5.**
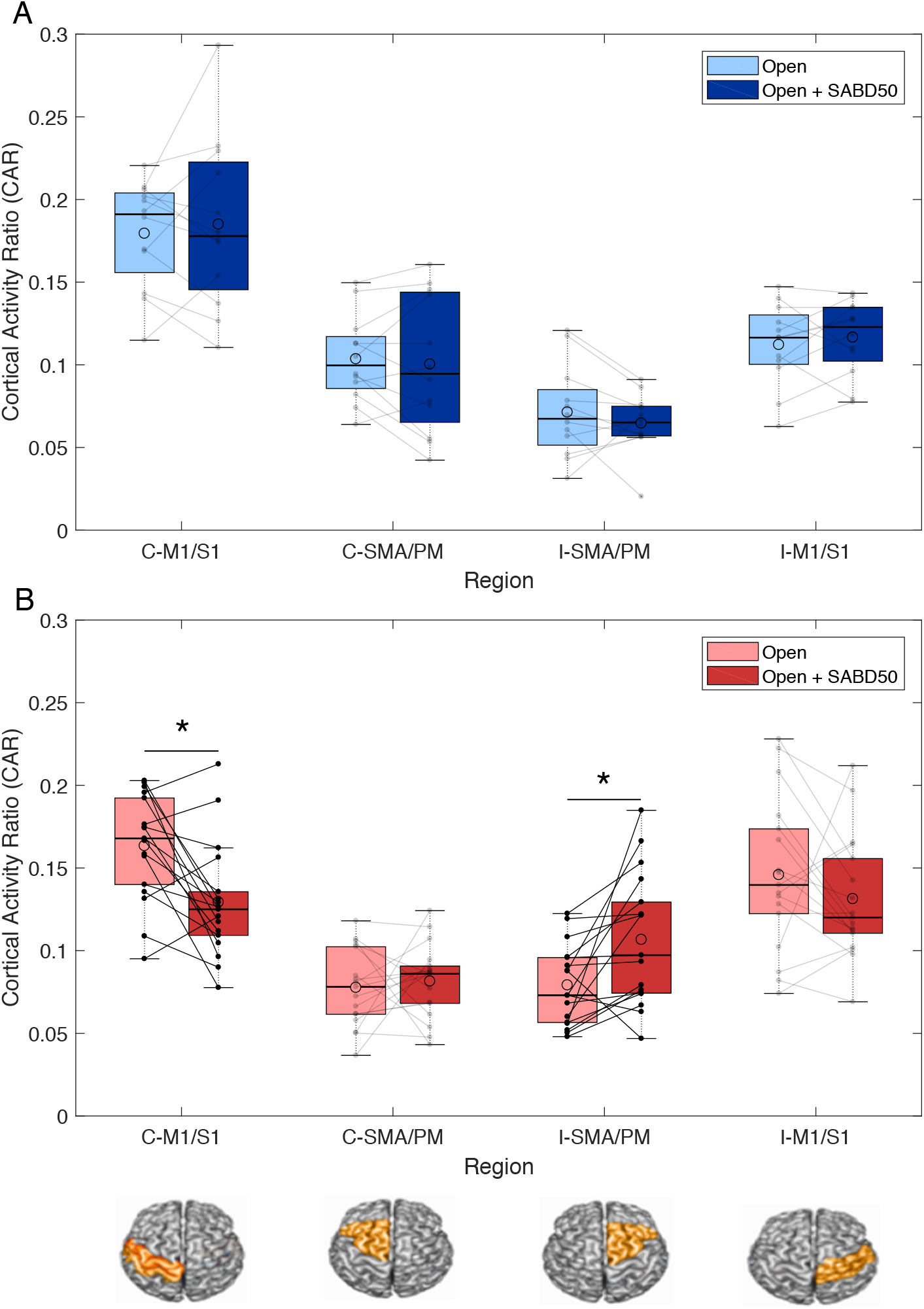
Cortical regions driving SABD-dependent reliance on the ipsilateral hemisphere. (A) Boxplots with individual data overlaid depicting cortical activity ratio (CAR) during hand opening (light blue) and hand opening while lifting against 50% max shoulder abduction (dark blue) across the 4 regions of interest in healthy controls. No changes in CAR are observed in any of the regions between the two tasks. (B) Boxplots with individual data overlaid depicting CAR during hand opening (light red) and hand opening while lifting against 50% max shoulder abduction (dark red) across the 4 regions of interest in individuals with stroke. Individuals demonstrated a decrease in activity in contralateral (ipsilesional) primary sensorimotor cortex (M1/S1) and an increase in ipsilateral (contralesional) secondary motor areas (SMA/PM) with the addition of SABD. ROIs are depicted below the figure. The median is shown by the horizontal black line and the mean is illustrated by the large open circle. C = contralateral, I = ipsilateral. * p < 0.05.

We further compared the between-task difference in the sum of absolute amplitude in i-SMA/PM and c-M1/S1, the 2 significant areas for CAR measure in individuals with stroke. Data were log transformed to normalize the data. Due to between-task differences in signal to noise ratio greater than 2 standard deviations from the mean difference thus making the comparison of absolute amplitude between conditions invalid, 2 participants were removed. Paired t-tests showed that the absolute amplitude of activity was increased in ipsilateral secondary motor areas with the addition of SABD (*t*(14) = 3.08, *p* = 0.008), but there was no difference between conditions for contralateral primary sensorimotor cortices (see Supplementary Figure 1).

### 3.4 Relationship Between Cortical Activity and Hand Opening Ability

Linear regression reported a positive correlation between involuntary grasping pressure and the CAR measure from i-SMA/PM during the Open + SABD50 condition (*R* = 0.65, *p* = 0.022; Figure 6A). Thus, individuals who showed more involuntary grasping forces when attempting to open during the SABD condition tended to show greater activity in ipsilateral secondary motor areas during that task. Meanwhile, there was no association between involuntary grasping pressure and activity in c-M1/S1 during this condition (*R* = −0.20, *p* = 0.53; Figure 6B).

**Figure 6.**
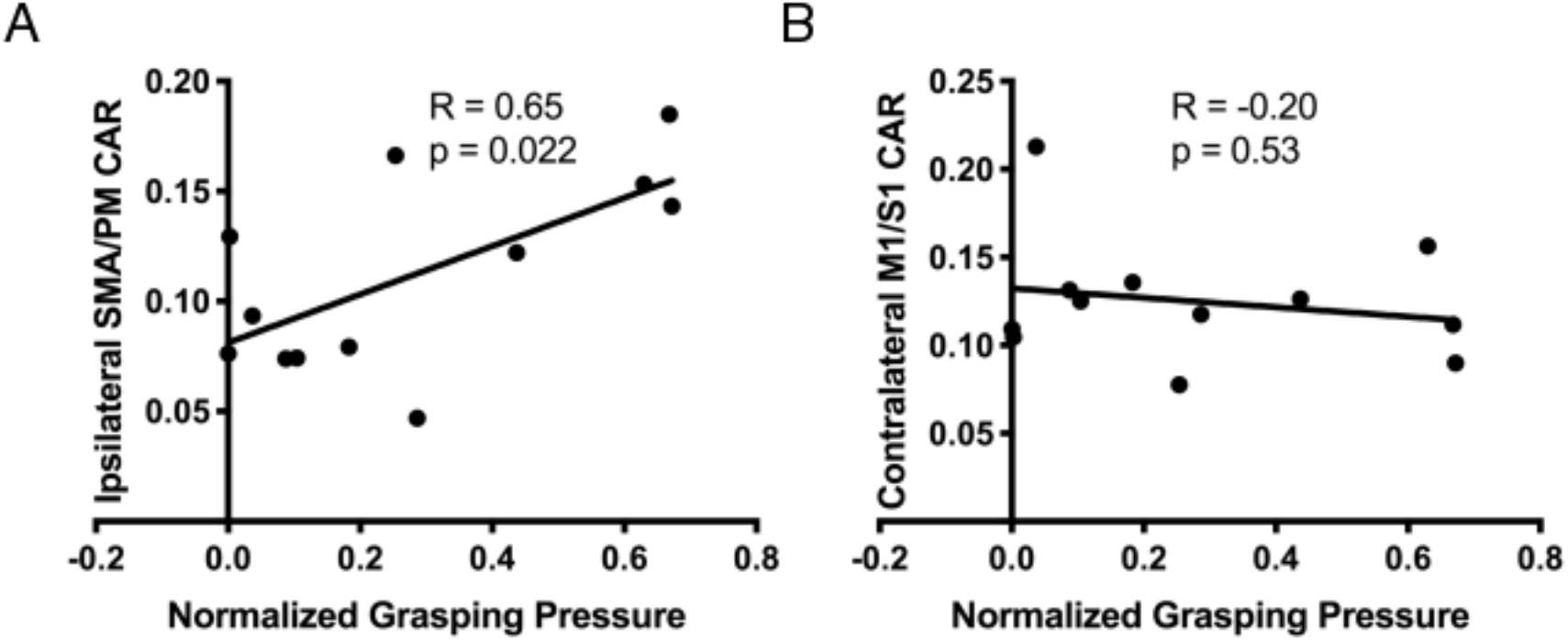
Association between cortical activity and hand opening ability in individuals with stroke. (A) Comparison of normalized grasping pressure during the hand opening + SABD50 condition and cortical activity ratio (CAR) in ipsilateral secondary motor areas (SMA/PM) during that task. Greater involuntary grasping pressure (i.e., reduced hand opening ability) is associated with greater activity in ipsilateral secondary motor areas in individuals with stroke. (B) Comparison of normalized grasping pressure during the hand opening + SABD50 condition and CAR in contralateral primary sensorimotor cortex (M1/S1) during that task. There is no association between activity in contralateral primary sensorimotor cortex and grasping pressure.

## IV. Discussion

We sought to evaluate the potential compensatory role of ipsilateral (i.e., contralesional) secondary motor regions post-stroke and their capacity to support hand function as compensation for damage to corticospinal tract. We found that individuals with stroke showed systemic changes in structural morphometry within ipsilateral secondary motor regions relative to the paretic arm in the form of increased gray matter density. Furthermore, when looking at cortical activity related to the hand, we found that the increased demand of SABD during attempted hand opening increased reliance on the i-SMA/PM in individuals with stroke, but not controls. Crucially, this reliance on ipsilateral secondary motor areas was associated with SABD-induced impairments in hand opening ability due to the flexion synergy. The combination of this structural and functional evidence points to increased compensatory reliance on ipsilateral secondary motor areas post moderate to severe stroke, but with limited capacity to support proper hand function due to involuntary recruitment of wrist/finger flexor muscles.

If individuals with stroke are indeed relying more on ipsilateral secondary motor areas as compensation for contralateral CST and corticobulbar damage, we would expect to see systemic changes in structural morphometry in these regions. This expectation is based on the known relationship between functional activity and both synaptogenesis and dendritic growth commonly seen in animal training models^33,34^. In line with these expectations, we saw increased GM density within ipsilateral secondary motor areas, specifically in the premotor cortex, in individuals with stroke compared to controls. Increases in GM density are proposed to indicate potential synaptogenesis, dendritic growth, or gliogenesis within these regions^34^. The observed changes may reflect a combination of a greater reliance on the non-paretic limb, associated with a high prevalence of learned non-use^5,35^ and a compensatory increased reliance on the ipsilateral projecting, cortico-bulbospinal tracts controlling the paretic limb in this population with more severe impairments. Recent evidence of an increase in medial reticulospinal structural integrity at the brainstem and cervical spinal cord at the ipsilateral side in individuals with hemiparetic stroke, compared to age matched controls, also fits this idea^6^. Findings in rodent models have similarly found increased dendritic growth and synapse proliferation in the ipsilateral cortex, particularly in animals showing excessive disuse^36,37^. Additionally, the more severe impairments prevalent in these individuals tends to lead to a reliance on compensatory strategies in everyday life, which has also been hypothesized to lead to associated structural changes in the ipsilateral cortex^38^.

Having established that individuals with chronic stroke demonstrate long-term changes in structure in i-SMA/PM, possibly indicating increased overall reliance on these regions, we then examined the functional capacity of these regions to support hand function. To this point, the majority of research on hand function post-stroke has focused on the role of CST damage and impairment levels^39,40^. Although damage to CST and accompanying corticobulbar tracts explains the presence of weakness post-stroke, it does not account for the loss of independent joint control such as the flexion synergy often observed in individuals with more severe impairments. The flexion synergy arises during arm lifting and reaching movements and leads to abnormal involuntary coactivation with elbow/wrist and finger flexors^20-23^. We hypothesize that this occurs because residual resources from remaining contralateral corticospinal and corticobulbar tracts become insufficient as the demand of the shoulder abduction increases, and consequently individuals rely more on uncrossed ipsilateral cortico-bulbospinal pathways, such as the corticoreticulospinal tract^6^, in compensation to carry out the motor task^41^. Unfortunately, although these ipsilateral pathways allow control of the shoulder, they reduce hand opening distally at the hand due to coactivation between shoulder abductors and wrist/finger flexors^23,26^. Our findings here support this hypothesis, as individuals with stroke demonstrated increased reliance on the ipsilateral hemisphere with the addition of an SABD load, along with reduced hand opening ability, whereas healthy controls, who have intact contralateral CST and corticofugal tracts, showed no effect of SABD.

We found that the observed shift to the ipsilateral hemisphere during the SABD task in the stroke group was driven by increased activity in i-SMA/PM as measured by CAR. This was confirmed when examining the overall absolute amplitude of cortical activity between the 2 conditions, suggesting that as the demand of the task increased, individuals with stroke attempted to use additional cortical resources from the ipsilateral secondary motor areas to execute the task. Ipsilateral secondary motor areas have been widely implicated for their compensatory role post-stroke, particularly in more impaired individuals. For instance, individuals post-stroke who used their paretic arm less in daily life, as measured by accelerometers, also showed greater activity in secondary motor areas during a grip task^42^. Similarly, increased secondary motor activity correlated with greater jerk in a reach to grasp movement, highlighting its compensatory role and inability to fully eliminate impairment^43^.

Importantly, SMA and PM serve as the primary origin for ipsilaterally-projecting cortico-bulbospinal tracts such as the corticoreticulospinal tract^8,11^. These tracts have been widely implicated in the presence of the flexion synergy due to their low-resolution output that spans multiple segments of the spinal cord and flexor bias in the ipsilateral arm^15^. Given that we see both increased activity in these areas and a correlation with reduced hand opening ability due to the flexion synergy during the SABD task, we argue this reflects increased recruitment of these ipsilateral cortico-bulbospinal pathways. This argument is grounded in work done in monkeys where lesions lead to an increase in strength of the corticoreticular projections^7^, and stimulation of reticulospinal pathways elicits activation of shoulder abductor and arm/hand flexor muscles^26^. It is unlikely that the increased activity in secondary motor areas during the SABD task in our study reflects use of descending projections from ipsilateral CST since these primarily originate from ipsilateral primary motor cortex, not SMA or PM, and these pathways do not sufficiently innervate the distal portions of the arm^7,44^. It is also important to note that we do not see a correlation between reduced activity in contralateral primary sensorimotor cortex and reduced hand opening ability during the SABD task, which corroborates the role of ipsilateral cortico-bulbospinal pathways originating primarily from secondary motor areas as the main initiator of the flexion synergy-related hand opening impairment.

The main difference observed here compared to monkey models of stroke is that dependence on ipsilateral (i.e., contralesional) secondary motor areas, and presumably ipsilateral corticobulbospinal tracts, does not appear to be sufficient for significant hand function recovery. Unlike humans, monkeys maintain the ability to still functionally use the hand following a pyramidal CST lesion, possibly due to a more viable rubrospinal tract innervating the hand.^2,45^. In fact, recovery of reaching and hand function correlates with increased structural connectivity within cortico-reticulospinal tract projections in monkeys^19,46^. However, the ability for these tracts to allow dexterous hand control seems limited in humans based on the current results, as well as recent findings showing that individuals with hemiparetic stroke show increased white matter integrity in the ipsilateral medial reticular spinal pathway at the brainstem and cervical spinal cord, which is correlated with their motor impairment severity^6^. This seems to contradict assertions that ipsilateral secondary motor areas may support recovery of hand function post-stroke^47,48^, at least in the case for hand opening. Instead, the current results fit better within the recently proposed framework by Li and colleagues in which increased reliance on the ipsilateral SMA/PM cortico-reticulospinal tract accounts for the movement impairments seen post-stroke^49^.

Interestingly, we also observed increased GM density within ipsilateral primary somatosensory cortex in addition to premotor cortex. One possibility is that this reflects reorganization within the sensory system to provide sensory information to motor outputs being generated by ipsilateral cortico-bulbospinal pathways. Indeed, preliminary evidence has shown that sensory information travels from the contralateral somatosensory cortex to the ipsilateral somatosensory cortex via the corpus callosum post-stroke^50^. Additionally, sensory recovery post-stroke has been associated with changes in both the contralateral and ipsilateral somatosensory cortex^51,52^. However, the majority of research on neural plasticity post-stroke has focused on the motor component of recovery, and thus it is difficult to prescribe the underlying neural mechanism driving this result.

### Clinical Implications

Our results in individuals with moderate to severe chronic stroke demonstrate the insufficient capacity, and actual further detriment, for ipsilateral secondary motor cortices to control hand opening. This finding points to the need to reengage the lesioned hemisphere in order to improve hand function. Although ipsilateral secondary motor areas allow sufficient control of the shoulder, they do not sufficiently innervate extensor muscles of the hand and instead lead to involuntary coactivation of flexor muscles. Therefore, they do not seem to offer a viable solution for basic hand function. It has been argued that perhaps in the case of individuals with severe impairment, increased reliance on ipsilateral-projecting corticobulbar pathways is the only option for staving off complete paralysis due to unsalvageable damage to the contralateral-projecting pathways from the lesioned hemisphere^53^. This is certainly possible, however, we have previously demonstrated the ability for individuals even with severe motor impairments to reengage the lesioned hemisphere and improve hand function following device-assisted training^54^. Considering it is also possible to reduce the flexion synergy through progressive SABD training^55,56^, future work targeting both the flexion synergy and finger/wrist extensor weakness may yield a solution towards improving both hand and upper extremity function via a reengagement of ipsilesional resources.

### Limitations

One of the main limitations of this study is the inability to directly measure activity within the corticoreticulospinal tract. We are limited to the cortex when using EEG to look at cortical activity related to the task, and thus cannot directly measure use of specific pathways. However, previous work has indeed shown structural changes in this pathway post-stroke, especially in individuals with more severe impairments^6,57^, supporting the notion of potential increased compensatory reliance. EEG also allows us to look at cortical activity during tasks involving SABD, which would be impractical inside an MRI scanner.

It is also worth noting that this experiment only looked at the effect of SABD on hand opening. Findings may be different for hand closing compared to opening. Ipsilateral corticoreticulospinal tract makes substantial innervations to the flexor muscles of the wrist and fingers, and thus may enable sufficient hand closing control, at least for power grasps^2,58^. This could explain why extensor weakness of the fingers is usually a more significant problem than flexor weakness post-hemiparetic stroke^22,59,60^.

## Data Availability

Data is available upon request.

## Acknowledgements

We would like to thank Dylan Fitzsimons for his assistance with EEG data collection.

